# First Point-of-Care Diagnostic Test for Beta-Thalassemia

**DOI:** 10.1101/2022.06.20.22276414

**Authors:** Ran An, Alireza Avanaki, Priyaleela Thota, Sai Nemade, Amrish Mehta, Umut A. Gurkan

**Affiliations:** Department of Mechanical and Aerospace Engineering, Case Western Reserve University, Cleveland, OH, USA; HemexHealth, Inc, Portland, OR, USA; Plasma Lab, Jalgaon, Maharashtra, India; Apple Diagnostics Lab in Ghatkopar, Mumbai, India; Department of Biomedical Engineering, Case Western Reserve University, Cleveland, OH, USA; Case Comprehensive Cancer Center, Case Western Reserve University, Cleveland, OH, USA

**Author notes:** **Corresponding authors:** Ran An, PhD, Senior Research Associate, Case Western Reserve University, Office: Glennan 611, 10900 Euclid Ave., Cleveland, OH 44106., Umut A. Gurkan, PhD, Warren E. Rupp Associate Professor, Case Western Reserve University, Office: Glennan 616B, 10900 Euclid Ave., Cleveland, OH 44106.

## Abstract

Hemoglobin (Hb) disorders are among the most common monogenic diseases affecting nearly 7% of the world’s population. Among various Hb disorders, approximately 1.5% of the world population carry *β*-thalassemia (*β*-Thal), affecting 40,000 newborns every year. Early screening and timely diagnosis are essential in *β*-thalassemia patients for prevention and management of later clinical complications. However, in Africa to Southern Europe, Middle East, and Southeast Asia, where *β*-thalassemia is most prevalent, diagnosis and screening of *β*-thalassemia is still challenging due to the cost and logistical burden of laboratory diagnostic tests. Here, we present Gazelle, a paper-based microchip electrophoresis platform, that enables the first point-of-care diagnostic test for *β*-thalassemia. We evaluated the accuracy of Gazelle for *β*-Thal screening in 372 subjects in the age range of 4 – 63 years at Apple Diagnostics labin Mubai, India. Additionally, 30 blood samples were prepared to mimic *β*-Thal intermediate and *β*-Thal major samples. The Gazelle detected levels of Hb A, Hb F, and Hb A_2_ demonstrated high correlations with the results reported by the laboratory gold standard, high performance liquid chromatography (HPLC) yielding a Pearson Correlation Coefficient = 0.99. The ability to obtain rapid and accurate results suggest that Gazelle can be suitable for large-scale screening and diagnosis of *β*-Thal.

## INTRODUCTION

Hemoglobin (Hb) disorders are among the world’s most common monogenic diseases [1]. *β*-thalassemia (*β*-Thal) is caused by single mutations resulting in small deletions or insertions within the *β-globin* gene or its immediate flanking sequence or gross deletions resulting in reduced production of normal hemoglobin (Hb A). Globally, approximately 1.5% of the world population carry *β*-Thal, affecting 40,000 newborns every year [2, 3]. It is estimated that over 90% of patients with *β*-Thal live in low-and-middle resource settings including Africa, the Middle East, Southeast Asia, and Southern Europe [4]. Successful screening and prevention programs have reduced the number of persons affected by *β*-Thal in countries such as Cyprus, Greece, and Italy [5]. However, such programs have not been widely implemented in other areas due in large part to the cost and logistical burden of laboratory diagnostic tests [5].

There are many *β*-Thal syndromes. The most common ones are: *β*-Thal major, *β*-Thal intermedia, and *β*-Thal trait. *β*-Thal major and *β*-Thal intermedia are characterized by an elevated fetal hemoglobin percentage (HbF%.) They are associated with severe moderate, or mild anemia requiring repeated transfusions resulting in a, a risk of iron overload. *β*-Thal trait is characterized by high HbA% along with increased levels of Hemoglobin A_2_ (HbA_2_% > 3.5%) and is associated with mild or no anemia and with variable microcytosis [6]. The current centralized tests used for screening and diagnosing *β*-Thal is high performance liquid chromatography (HPLC). These tests rely on often unaffordable (15k - 35k US Dollar, or 90k – 210k Ghanaian Cedi) specialized instruments, state-of-the-art laboratory facilities, and highly trained personnel, which are lacking in low resource settings where *β*-Thal is most prevalent [2]. As a result, there is a need for affordable, portable, easy-to-use, accurate, point-of-care (POC) tests to facilitate decentralized *β*-Thal testing in low-resource settings.

Several POC diagnostic systems for several hemoglobin variants such as Hb S have been described [7-9] based on testing methods such as the sickle cell solubility test and antibody-based lateral flow assays such as Sickle SCAN™ and HemoType SC™ [10-12]. However, there is currently no POC test available for *β*-Thal detection. In a 2019 report, the World Health Organization (WHO) listed hemoglobin testing as one of the most essential in vitro diagnostic (IVD) tests for primary care use in low and middle income countries [13]. Furthermore, hemoglobin electrophoresis has recently been added to the WHO essential list of IVDs for diagnosing sickle cell disease (SCD) and sickle cell trait [14]. Leveraging the WHO recognized Hb electrophoresis test, we developed a paper-based, miniaturized Hb electrophoresis platform, Gazelle™ (**Fig.1**) [15-22]. Gazelle has been tested in clinical studies in 4 different countries with more than 700 subjects, and demonstrated capability of identifying major Hb variants including HbA, HbE, HbS, and HbF, in adults as well as in newborns with SCD, sickle cell trait, Hemoglobin C disorder, and Hemoglobin E Disorder [15-22].

**Figure 1.**
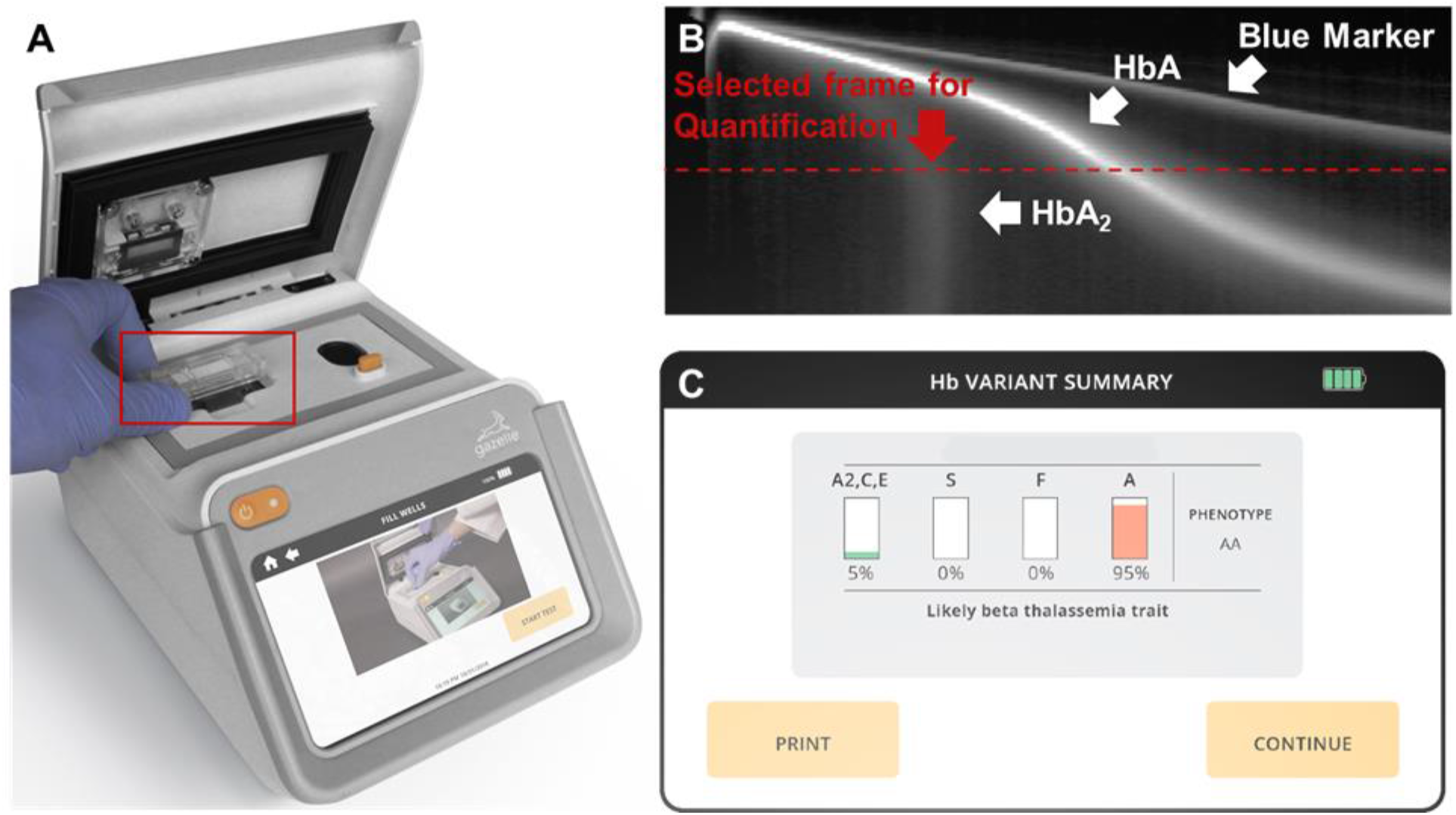
Gazelle for screening beta-thalassemia. **(A)** Gazelle paper-based microchip electrophoresis using a disposable cartridge **(red box)** at the point of need. **(B)** Applying an internally integrated data analysis algorithm, the generated space-time plots based on the captured images are used for identification and quantification Hb variants in real time. **(C)** At the end of each test, Gazelle algorithm automatically reports the identified and quantified Hb variant results, and the patient phenotype.

Here, we implemented a customized image analysis algorithm for accurate quantification of HbA_2_ for the first time, in addition to HbA, HbF, and Hb C/E. Additionally, we describe a test for evaluating the diagnostic performance of this platform for identifying *β*-Thal major, *β*-Thal intermedia, and *β*-Thal trait over 372 subjects in the age range of 4 – 63 years at Apple Diagnostics lab in Ghatkopar, Mumbai, India. Additionally, 32 blood samples were prepared to mimic *β*-thalassemia intermediate and *β*-thalassemia major samples. The Gazelle detected levels of Hb A, Hb F, and Hb A_2_ demonstrated high correlations with the results reported by laboratory gold standard high performance liquid chromatography (HPLC) yielding a Pearson Correlation Coefficient = 0.99. These results suggest that Gazelle-Multispectral is potentially suitable for large-scale *β*-Thal testing.

## METHODS

### Study Design and Oversight

We conducted a prospective diagnostic accuracy study on Gazelle for detecting Hb variants including HbA, Hb, and HbA_2_ at Apple Diagnostics lab in Ghatkopar, Mumbai, India, under an IRB-approved study protocol [IEC@IISS (International Institute of Sleep Sciences IRB number); Reg No.: ECR/177/Indt/MH-2014/RR-19]. The results obtained from Gazelle were compared to the results reported by the reference (“Gold-standard”) tests using HPLC in India. All samples have been tested within 7 days of collection. All authors have reviewed and analyzed the data and attest to their accuracy and completeness as well the fidelity of adherence to the study protocol.

### Study Populations and Procedures

This test was conducted at Apple Diagnostics in Mumbai, India. Blood samples were collected from the subjects at screening camps and education and awareness programs conducted by Plasma Lab in Jalgaon district, Maharashtra, India. All these subjects were consented to participate in the study. Subjects were excluded only if there had been a blood transfusion in the preceding 3 months or if they were unable to give informed consent. The study was approved by the IRB of the International Institute of Sleep Sciences (IISS) and informed consent obtained from each participant’s parent or guardian if required. All the blood samples were collected on site into EDTA blood collection tubes and stored at 4C until they were tested with Gazelle. Blood samples were tested within one week of collection. The left-over blood was stored at 4C for HPLC testing using a D-10 HPLC system (Bio-Rad Laboratories, Hercules, CA, USA). The laboratory technician who conducted the tests on Gazelle had basic laboratory skills such as pipetting and vortexing and was able to independently perform the tests with less than 2 hours training.

### Gazelle Test Procedure

The technicians performed the tests according to the Gazelle-Multispectral instructions for use as published previously [21]. Similarly, all buffer compositions have been published previously [21]. At the concousion of the 8 minute test, Gazelle automatically reports the percentages of each hemoglobin type present in the blood sample as well as an interpretative statement.

### Confirmatory Laboratory Procedures

Samples that were stored at 4 °C degrees were retrieved. 5 microlitres of sample was pipetted and diluted with 1500 microliters of distilled water. Diluted hemolysates were arranged on racks and loaded into Bio-Rad D-10. Bar coded Samples IDs were then edited to match sample IDs. Each sample was run for approximately 6 mins.

### Gazelle-Multispectral Data Analysis

A customized data analysis algorithm was integrated in the Gazelle system. This data analysis algorithm automatically identifies β-Thal major, β-Thal intermedia, and β-Thal trait based on Hb band migration pattern as described previously [21, 23]. The data analysis algorithm also automatically quantifies the relative percentages of HbA, HbF, and HbA_2_ in addition to other hemloglobins as previously reported. Gazelle’s reported Hb variant identification and quantification results were compared with the ones reported by HPLC using Pearson correlation and Bland-Altman analysis. Gazelle-Multispectral sensitivity, specificity in identification of β-Thal major, β-Thal intermedia, and β-Thal trait were calculated for the study population compare to HPLC reported results.

## RESULTS

### Test Population and Result Reporting

In this study, we have conducted clinical testing over 372 subjects in the age range of 4 – 63 years at Apple Diagnostics lab in Ghatkopar, Mumbai, India, under local IRB-approved study protocol. Additionally, 32 blood samples were artificially prepared to mimic *β*-Thal major, and *β*-Thal intermedia samples.

The Gazelle algorithm verifies the quality of test results according to the internally embedded data quality control (QC) method. According to the QC method, Gazelle-Multispectral organizes test results under one of the 3 categories: 1) ‘Valid’ test; 2) ‘Uninterpretable’ test; and 3) ‘Inconclusive’ test. The ‘Valid’, ‘Uninterpretable’, and ‘Inconclusive’ tests were defined according to published recommendations in previous publication [24] and the STARD guidelines [25]. If a test is performed as expected according to objective standards, the data analysis algorithm recognizes the test as a ‘Valid’ test and reports the test result. If a test includes poor migration of the blue control marker, electrical connectivity issue, or faculty cartridges, the data analysis algorithm recognizes the test as an ‘Uninterpretable’ test. If a test is performed adequately according to an objective set of standards, but has a quantification confidence value that is lower than the preset threshold value, then the data analysis algorithm recognizes the test as an ‘Inconclusive’ test. Reasons for ‘Inconclusive’ tests include appearance of a Hb variant band or bands at or close to the borderline region between two adjacent detection windows. In this study, 394 out of 404 (97.5%) tests were recognized as ‘Valid’, while 10 out of 404 (2.5%) tests were recognized as ‘Inconclusive’.

### Gazelle Result Interpretation Criteria

The Gazelle result interpretation criteria was established based upon clinical recommendations from key opinion leaders and using a large training dataset collected from field studies. The Gazelle *β*-Thal algorithm defines *β*-Thal major, *β*-Thal intermedia, and *β*-Thal trait based on the detected HbA, HbF, and HbA_2_ levels. The algorithm reads and interprets the data in the following sequences: 1) first the algorithm checks if HbF ≥ 60%, and HbA < 40% with no other hemoglobins over 10%, if true the algorithm calls the sample *β*-Thal major or *β*-Thal intermedia. 2) when HbF < 60% and HbA ≥ 40% with no other hemoglobins over 10%, the algorithm measures the HbA_2_ level and if HbA_2_ > 3.5%, then the algorithm calls the sample as *β*-Thal trait.

### Gazelle-Multispectral Hb variant quantification demonstrated high correlation with HPLC

Pearson correlation analysis and Bland-Altman analysis were performed on the 394 tests recognized as ‘Valid’. The correlation plots include the Gazelle determined Hb variant levels (*y* axis) versus the Hb variant levels reported by the HPLC (*x* axis) including HbA (black), HbF (orange), HbA_2_ (red, **Fig. 2A**). The Bland-Altman analysis includes the difference between Gazelle quantified levels of HbA, HbF, and HbA_2_ and the ones determined by HPLC (*y* axis) over the entire range of Hb levels detected (*x* axis, **Fig. 2B**). The result from the Pearson correlation analysis were a Pearson correlation coefficient (PCC) of 0.99 for all three Hb variants. Bland-Altman analysis results indicates Gazelle determines blood Hb variant levels with mean bias of +1.2%, with upper and lower limit of agreement at 4.8% and −2.4%, respectively. Together, these results demonstrated good agreement between the Hb variant levels determined by Gazelle and the ones reported from HPLC (**Fig. 2**).

**Figure 2.**
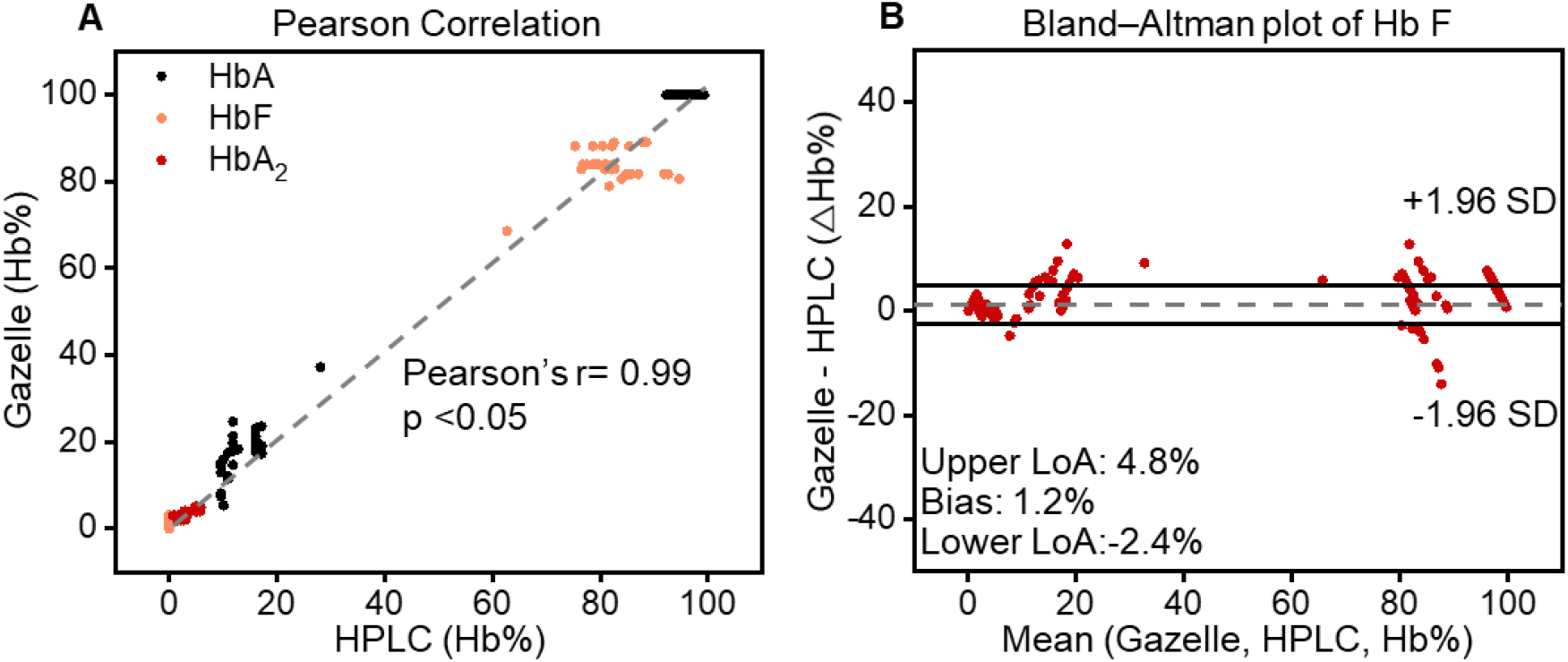
Gazelle accurately identifies and quantifies HbA, HbF, and HbA_2_. **(A)** Pearson correlation showed Gazelle identified and quantified HbA (black), HbF (orange), and HbA_2_ (red) demonstrated strong correlation with the results reported by laboratory standard test HPLC at Person coefficient correlation (PCC) = 0.99, p < 0.05). **(B)** Bland-Altman analysis demonstrated that Gazelle tests had an overall mean bias of 1.2% (doted gray line) with upper and lower limit of agreement (LoA) at 4.8% and −2.4%, respectively).

### Sensitivity and specificity of Gazelle *β*-Thalassemia Testing

In this clinical study, Gazelle test results included disease (*β*-Thal major and *β*-Thal intermedia), trait (*β*-Thal trait) and normal (categorized using data interpretation criteria described above). Comparing the results reported from the laboratory standard test HPLC, Gazelle identified subjects with disease, *β*-Thal major and *β*-Thal intermedia, from normal subjects and subjects with the *β*-Thal trait with 100% sensitivity and specificity (**Table 1**), 6 normal subjects were identified as *β*-Thal trait. Sensitivity and specificity for identifying subjects with *β*-Thal trait from normal subjects (Trait vs. Normal) were 100% and 98.3%, respectively.

**Table 1:**
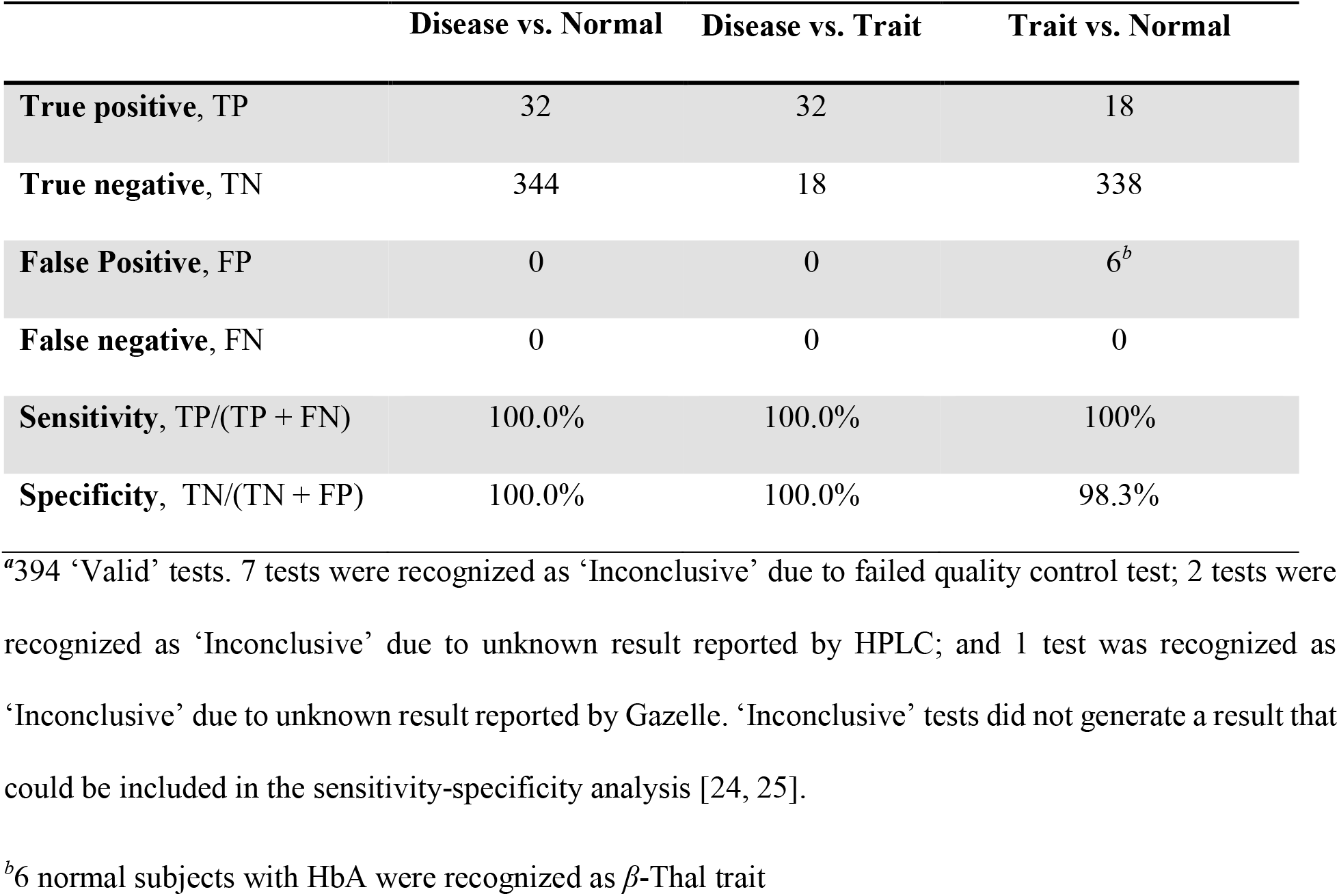
Gazelle-Multispectral screening sensitivity, specificity, positive predictive value (PPV), and negative predictive value (NPV) in comparison to reference standard method^*a*^.

## Conclusions

*β*-Thal major and *β*-Thal intermedia are characterized by an elevated fetal hemoglobin percentage (HbF%) and *β*-Thal trait is characterized by high HbA% with increased levels of HbA_2_% [6]. Proper and timely management of *β*-Thal relies on early and accurate detection. Quantification of HbA and key Hb variants including HbF and Hb HbA_2_ are essential for accurate detection of *β*-Thal. The previous Gazelle system has demonstrated its utility in detecting anemia [23] and hemoglobinopathies including SCD, sickle cell trait, Hemoglobin C disorder, and Hemoglobin E Disorder [15-22]. Here, we report the updated version Gazelle which enables, for the first time, POC detection of *β*-Thal.

In this clinical study conducted among 404 subjects in Ghatkopar, Mumbai, India, Gazelle demonstrated 100% sensitivity and specificity for identifying *β*-Thal major/intermedia vs. healthy subjects; and subjects with *β*-Thal major/intermedia vs. *β*-Thal trait. Additionally, Gazelle demonstrated 100% sensitivity and 98.3% specificity for identifying *β*-Thal trait vs. healthy subjects. Recommended practice in Hb testing is that all positive test results are confirmed with a secondary method prior to final diagnostic decision making and treatment initiation [26]. Therefore, all disease positive tests would likely result in a secondary confirmatory test that should eliminate potential false positives if there is any.

In summary, Gazelle provides, for the first time, affordable and rapid POC solution for detecting *β*-Thal. The Gazelle reader provide 1) animated on-screen instructions with step-by-step guidance for test operation procedures to minimize user errors; and 2) a data analysis algorithm that automatically reports Hb variant levels and predicted disease status. Overall, the results reported in this study suggest that Gazelle is potentially suitable for large-scale *β*-Thal testing in low resource regions where the prevalence of *β*-Thal is high.

## Data Availability

All data produced in the present study are available upon reasonable request to the authors

## ACKNOWLEDGEMENTS

Authors acknowledge National Heart Lung and Blood Institute Small Business Innovation Research Program (R44HL140739, R41HL151015), National Heart Lung and Blood and the National Center for Complementary & Integrative Health (NCCIH) 1U54HL143541, National Institute of Diabetes and Digestive and Kidney Diseases Small Business Innovation Research Program (R41DK119048), and NHLBI R01HL133574 and T32HL134622. This article’s contents are solely the responsibility of the authors and do not necessarily represent the official views of the National Institutes of Health.

## AUTHOR CONTRIBUTIONS

RA and AA contributed to the proof-of-concept experiments and initial development. PT, SN, and AM helped with the planning and execution of clinical testing, including human subject research protocol development, subject recruitment, blood sample collection, and testing. RA, and AA performed the data analysis, prepared the tables, figures, figure captions, and supplementary information. RA drafted the manuscript. RA, AA, PT, SN, AM, and UAG reviewed and edited the manuscript.

## DATA AND MATERIALS AVAILABILITY

All reasonable requests for materials and data will be fulfilled by the corresponding author of this publication.

